# Voxel-level Classification of Prostate Cancer Using a Four-Compartment Restriction Spectrum Imaging Model

**DOI:** 10.1101/2020.07.25.20162172

**Authors:** Christine H Feng, Christopher C Conlin, Kanha Batra, Ana E Rodríguez-Soto, Roshan Karunamuni, Aaron Simon, Joshua Kuperman, Rebecca Rakow-Penner, Michael E Hahn, Anders M Dale, Tyler M Seibert

**Author notes:** Corresponding Author: Tyler Seibert, MD, PhD, Altman Clinical and Translational Research Institute, 9500 Gilman Dr. #0861, La Jolla, CA 92093.

## Abstract

**Purpose:** Diffusion MRI is integral to detection of prostate cancer (PCa), but conventional ADC cannot capture the complexity of prostate tissues. A four-compartment restriction spectrum imaging (RSI_4_) model was recently found to optimally characterize pelvic diffusion signals, and the model coefficient for the slowest diffusion compartment, RSI_4_-C_1_, yielded greatest tumor conspicuity. In this study, RSI_4_-C_1_ was evaluated as a quantitative voxel-level classifier of PCa.

**Methods:** This was a retrospective analysis of 46 men who underwent an expanded-acquisition pelvic MRI for suspected PCa. Twenty-three men had no detectable cancer on biopsy or clinical follow-up; the other 23 had biopsy-proven PCa corresponding to a lesion on MRI (PI-RADS category 3-5). High-confidence cancer voxels were delineated by expert consensus, using imaging data and biopsy results. The entire prostate was considered benign in patients with no detectable cancer. Diffusion images were used to calculate RSI_4_-C_1_ and conventional ADC. Voxel-level discrimination of PCa from benign prostate tissue was assessed via receiver operating characteristic (ROC) curves generated by bootstrapping with patient-level case resampling. Specifically, we compared RSI_4_-C_1_ and conventional ADC on mean (and 95% CI) for two metrics: area under the curve (AUC) and false-positive rate for a sensitivity of 90% (FPR_90_). Classifier images were also compared.

**Results:** RSI_4_-C_1_ outperformed conventional ADC, with greater AUC [0.977 (0.951-0.991) vs. 0.921 (0.873-0.949)] and lower FPR_90_ [0.033 (0.009-0.083) vs. 0.201 (0.131-0.300)].

**Conclusion:** RSI_4_-C_1_ yielded a quantitative, voxel-level classifier of PCa that was superior to conventional ADC. RSI classifier images with a low false-positive rate might improve PCa detection.

## Introduction

Prostate cancer is the second most frequent malignancy in men worldwide and is a common cause of cancer deaths in men^1^. Strategies to improve outcomes for men with prostate cancer seek to optimize detection, staging, and clinical risk stratification. The 12-core systematic biopsy remains a common method for initial diagnosis and Gleason grading of prostate cancer, but is prone to sampling errors that can drastically influence risk stratification and treatment^2,3^. Multiparametric magnetic resonance imaging (MRI) has become increasingly popular for its added value in identifying suspicious lesions for targeted biopsy^4–6^.

Clinical multiparametric MRI currently includes diffusion-weighted imaging (DWI) and apparent diffusion coefficient (ADC) maps to determine a qualitative risk of clinically significant cancer (PI-RADS v2^7^). However, conventional ADC is a measurement of overall diffusion rate of water within a voxel and can be influenced by multiple factors. It has shown correlation with presence of malignancy, but remains limited by motion sensitivity^8^, magnetic field inhomogeneity^9^, and high false-positive rates from inflammation, hemorrhage, or benign lesions that limit tumor conspicuity and localization^10–12^. Twenty-eight percent of PI-RADS (v2) category 5 lesions (the highest level of suspicion) do not yield a diagnosis of clinically significant cancer^13^, and false positive rates are even higher for category 3 and 4 lesions.

Advanced diffusion models use additional parameters to separate and characterize diffusion signals originating from various microstructural compartments within a voxel^14–16^. Restriction spectrum imaging (RSI) is a flexible framework that allows for a mixture of restricted intracellular, hindered extracellular, and freely diffusing water compartments to be probed with clinically relevant protocols^10,17^. The RSI technique models signal intensity as a function of *b*-value using a series of exponential decay functions, each representing a diffusion compartment with a specific, pre-determined ADC^10,17^. Optimal compartmental ADCs were recently estimated for the prostate (and seminal vesicles) using RSI models of two to five tissue compartments^18^. The overall diffusion signal was better characterized in models using more compartments, with the four-compartment model emerging as the best option by relative Bayesian information criterion^18^. In this study, we apply the four-compartment RSI model to the prostate and assess voxel-level accuracy for detection of prostate cancer, as compared to ADC.

## Methods

### Study Population

This was a retrospective study of 46 consecutive men who underwent screening pelvic MRI for suspected prostate cancer between August and December 2016 using an expanded acquisition protocol on a single scanner. Standard-of-care evaluations determined that 23 men had no detectable cancer, while another 23 men had prostate cancer attributable to a PI-RADS v2.1 category 3-5 lesion on MRI. This study was reviewed and approved by the UC San Diego Institutional Review Board (IRB #191878).

### MRI data acquisition and post-processing

Scans were collected on a 3T clinical MRI scanner (Discovery MR750, GE Healthcare, Waukesha, WI, USA) using a 32-channel phased-array body coil centered on the pelvis. Each patient underwent a high-resolution, T2-weighted sequence with identical scan coverage as the multi-shell DWI volume (TR: 6225 ms, TE: 100 ms, resolution: 0.39×0.39 mm, matrix: 512×512, slice thickness: 3 mm). A multi-shell diffusion volume was also acquired for each patient, sampling 5 *b*-values (0, 200, 1000, 2000, and 3000 s/mm^2^) at 6 unique gradient directions (TR: 5000 ms, TE: 80 ms, resolution: 1.6×1.6 mm, matrix: 128×128, slice thickness: 3 mm). The *b* = 0 s/mm^2^ volumes were acquired using forward and reverse phase encoding to allow for correction of B0-inhomogeneity distortions. The acquisition time for the diffusion volume was approximately 5 minutes.

Post-processing of MRI data was completed using in-house programs written in MATLAB (MathWorks, Natick, MA, USA). Diffusion data were corrected for distortions arising from B0 inhomogeneity, gradient nonlinearity, and eddy currents^10,19^. Conventional ADC was calculated for each voxel using distortion-corrected DWI sequences performed with *b*-values of 0, 200, and 1000 s/mm^2^.

### Prostate Data Extraction

The body, prostate, and prostate cancer lesion regions of interest (ROIs) were contoured by a radiation oncologist using MIM (MIM Software Inc, Cleveland, OH, USA). Prostate cancer ROIs were defined directly on DWI volumes, using all available clinical information. Defining ROIs on DWI prevents inadvertent inclusion of benign tissue into the ROI due to subtle registration errors. ROIs were verified via consensus by two board-certified sub-specialist radiologists, M.H. and R.R.P. The finalized ROIs were exported as binary masks into a MATLAB-compatible format that matched the resolution of the DWI volumes.

### RSI Models of Prostate Diffusion

The relationship between corrected signal intensity and b-value was modeled as a linear combination of exponential decays, where *S*_*corr*_(*b)* represents the noise-corrected DWI signal at a particular *b* value, *C* represents signal contribution of each compartment to the overall signal, and *D* represents the estimated ADC value for that compartment.

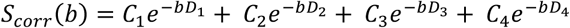

Noise correction for DWI volumes was performed as previously described^18^. Signal intensity was normalized by the median *b*=0 intensity within the prostate for each subject. Optimal *D* values for each compartment were previously determined by fitting the multi-shell DWI data from all voxels within the benign body and prostate cancer lesion ROIs^18^. The compartments are ordered from lowest to highest *D*, with the first compartment of each model describing the most restricted mode of diffusion. Prior work identified the four-compartment RSI model as optimally describing the diffusion signal from the prostate and prostate cancer^18^. For this model, the optimal ADCs for the compartments were 1.0 e-4, 1.8 e-3, 3.6 e-3, and >>3.0 e-3 mm^2^/s, approximately representing restricted, hindered, free diffusion, and flow, respectively^18^.

### Classification of Benign Prostate Tissue and Prostate Cancer

Prostate cancer conspicuity was related to the compartment with slowest diffusion in each model, called C_1_, with increased cancer conspicuity for the four-compartment model. Here, C_1_ for the four-compartment RSI model (RSI_4_-C_1_) was assessed for its ability to correctly identify benign prostate tissue and prostate cancer at the voxel level. Results with RSI were compared to those using standard ADC.

Classification of cancer and benign prostate voxels was assessed via 1000 bootstrap samples with case resampling at the patient level to yield means and 95% confidence intervals for performance metrics. Benign subjects contributed voxels from the entire prostate, and cancer subjects contributed voxels from only the high-confidence cancer ROIs. Voxels outside the high-confidence ROIs in patients with known cancer were excluded from statistical analysis because prostate cancer is notoriously multifocal and voxel-level ground-truth histopathology was not available. Area under the curve (AUC) of receiver operating characteristic (ROC) curves was the primary metric. Additionally, as a significant limitation of conventional ADC is poor specificity, performance was also assessed using the false positive rate for a sensitivity of 90% (FPR_90_). Statistical significance was assessed using paired t-tests with two-sided *α* = 0.05. P-values were truncated at p<10^−16^, if applicable.

Voxel-wise classifier maps were created by the logistic regression of RSI_4_-C_1_ (RSI_4_-C_1_ classifier) using all subjects. These maps were saved in DICOM format and overlaid on the T2 volume for visualization using MIM (MIM Software Inc, Cleveland, OH, USA) to indicate degree of suspicion for prostate cancer. ADC maps were generated for visual comparison.

## Results

Patient characteristics for cases with prostate cancer are in **Table 1**. Of the men with benign prostates on biopsy and/or surgical pathology, ten had PI-RADS category 1 prostates, two had PI-RADS category 2 lesions, eight had PI-RADS category 3 lesions, two had PI-RADS category 4 lesions, and one had a PI-RADS category 5 lesion.

**Table 1.** Characteristics of cases with prostate cancer. Clinical risk groups were designated per NCCN guidelines^34^ for favorable intermediate risk (FIR), unfavorable intermediate risk (UIR), high risk (HR), and very high risk (VHR).

| Case | Age | PI-RADS v2 | Lesion Location | PSA (ng/mL) | Gleason Grade Group | Pathology Specimen Type | Stage | Clinical Risk Group |
| --- | --- | --- | --- | --- | --- | --- | --- | --- |
| 1 | 59 | 5 | Right Transitional & Peripheral Zones | 7.77 | 2 | Systematic Biopsy | cT1cN0 | FIR |
| 2 | 54 | 5 | Right Peripheral Zone | 10.6 | 3 | Systematic Biopsy, Radical Prostatectomy | pT3aN0 | UIR |
| 3 | 71 | 5 | Right Peripheral & Transitional Zones | 4.04 | 5 | Systematic and Targeted Biopsy, Radical Prostatectomy | pT3aN0 | HR |
| 4 | 72 | 5 | Right Peripheral Zone | 5.7 | 2 | Systematic Biopsy | cT2bN0 | UIR |
| 5 | 54 | 5 | Right Peripheral Zone | 7.3 | 3 | Systematic Biopsy, Radical Prostatectomy | pT3aN0 | HR |
| 6 | 63 | 5 | Right Peripheral Zone | 16.83 | 3 | Systematic Biopsy, Radical Prostatectomy | pT3aN0 | HR |
| 7 | 74 | 5 | Right Peripheral Zone | 29.3 | 5 | Systematic Biopsy | cT2cN0 | HR |
| 8 | 53 | 4 | Left Peripheral Zone | 8 | 1 | Systematic Biopsy, Radical Prostatectomy | pT2cN0 | FIR |
| 9 | 66 | 4 | Right Peripheral Zone | 8.66 | 3 | Systematic and Targeted Biopsy | cT1cN0 | UIR |
| 10 | 67 | 5 | Left Peripheral Zone | 4.6 | 2 | Systematic and Targeted Biopsy | cT1cN0 | FIR |
| 11 | 62 | 5 | Left Peripheral Zone | 13.97 | 3 | Systematic Biopsy, Radical Prostatectomy | pT3aN0 | HR |
| 12 | 74 | 4 | Left Peripheral Zone | 4.4 | 3 | Systematic and Targeted Biopsy, Radical Prostatectomy | pT2aN0 | UIR |
| 13 | 50 | 3 | Left Peripheral Zone | 4.3 | 2 | Systematic Biopsy, Radical Prostatectomy | pT3aN0 | HR |
| 14 | 65 | 4 | Right Transitional Zone | 8 | 1 | Systematic Biopsy | cT1cN0 | LR |
| 15 | 81 | 5 | Anterior Transitional Zone | 8.5 | 2 | Systematic and Targeted Biopsy | cT2aN0 | FIR |
| 16 | 77 | 5 | Right Peripheral Zone | 3.47 | 4 | Biopsy (Systematic), Radical Prostatectomy | ypT2aN0 | HR |
| 17 | 70 | 4 | Left Peripheral Zone | 7.43 | 2 | Systematic and Targeted Biopsy | cT1cN0 | FIR |
| 18 | 58 | 4 | Left Peripheral Zone | 7.42 | 2 | Systematic and Targeted Biopsy, Radical Prostatectomy | mpT2cN0 | UIR |
| 19 | 62 | 4 | Left Peripheral Zone | 5 | 1 | Systematic Biopsy | cT1cN0 | LR |
| 20 | 68 | 4 | Right Peripheral Zone | 5.9 | 5 | Systematic and Targeted Biopsy, Radical Prostatectomy | mpT2cN0 | HR |
| 21 | 64 | 5 | Midline to Right Transitional Zone | 8.63 | 2 | Systematic and Targeted Biopsy | cT1cN0 | FIR |
| 22 | 51 | 5 | Diffuse Peripheral Zone | 33 | 5 | Systematic Biopsy | ypT3bN0 | VHR |
| 23 | 51 | 3 | Right Transitional Zone | 5.42 | 3 | Systematic Biopsy, Radical Prostatectomy | pT2bN0 | UIR |

RSI_4_-C_1_ outperformed conventional ADC as a quantitative, voxel-level classifier. RSI_4_-C_1_ had a greater AUC: mean 0.977 (95% CI 0.951-0.991), compared to 0.921 (0.873-0.949) for ADC (**Figure 1A**). The false positive rate was also lower for RSI_4_-C_1_: mean 0.033 (0.009-0.083), compared to 0.201 (0.131-0.300) for ADC (**Figure 1B**). Paired t-tests confirmed statistically significant differences in AUC and FPR_90_ between RSI_4_-C_1_ and conventional ADC (p<10^−16^, t=84.00 for AUC and p<10^−16^, t=-118.07 for FPR_90_). ROC curves for RSI_4_-C_1_ and ADC are presented in **Figure 2**, and demonstrate the improvement in false positive rate while maintaining high sensitivity. The distribution of benign and cancer voxels by normalized signal intensity of RSI_4_-C_1_ showed less overlap between the two groups of voxels compared to that of ADC (**Figure 3**). RSI_4_-C_1_ classifier output images and conventional ADC maps for representative subjects are shown in **Figure 4**.

**Figure 1.**
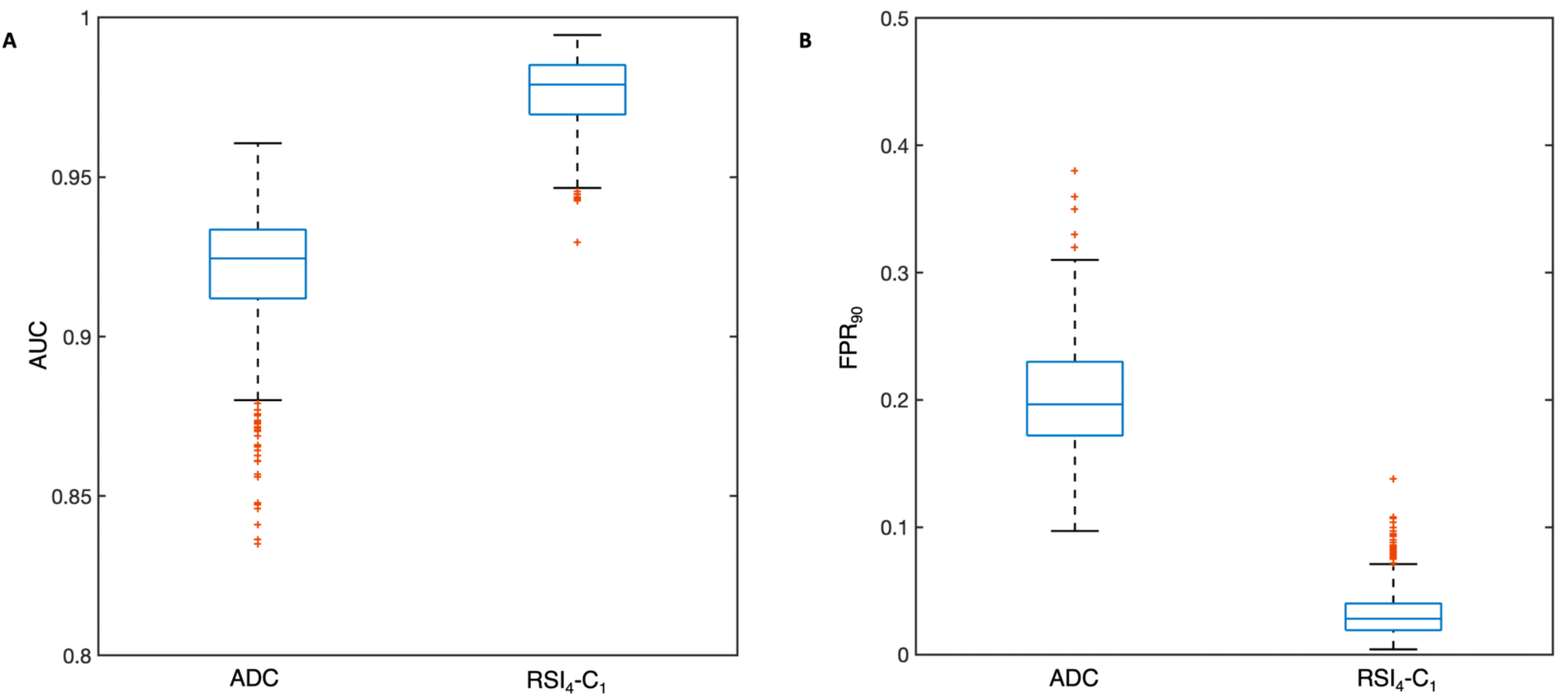
Box plots depicting distribution of performance metrics for 1000 patient-level bootstrap samples for A) area under the curve (AUC) and B) the false positive rate at 90% sensitivity (FPR_90_) for conventional ADC and RSI_4_-C_1_. Whiskers represent values within 1.5 times the interquartile range (IQR).

**Figure 2.**
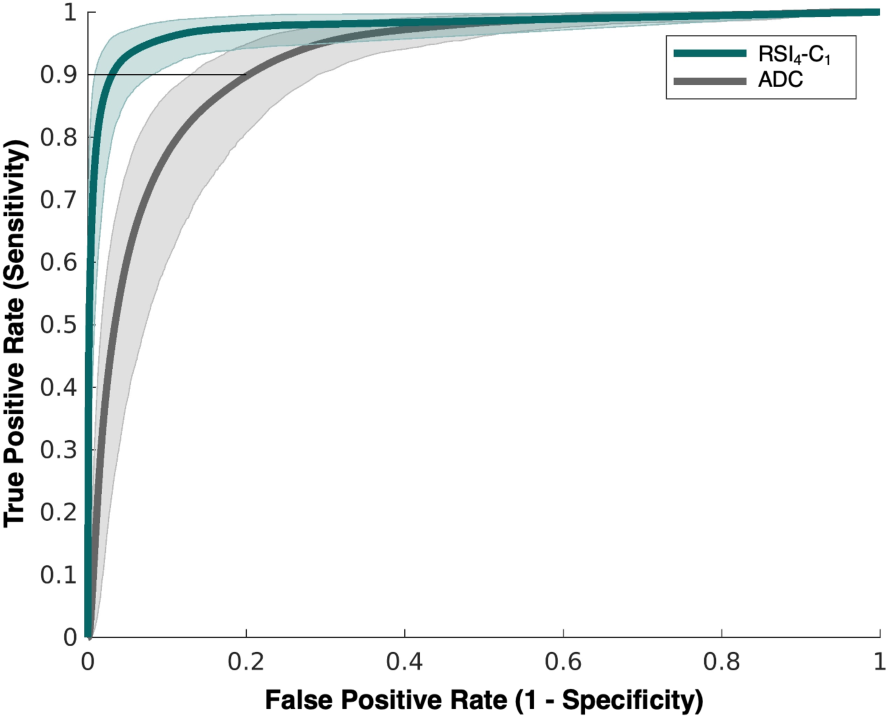
Receiver operating characteristic (ROC) curves for conventional ADC (grey) and RSI_4_-C_1_ (green) with confidence intervals indicated by shaded areas. FPR_90_ is highlighted by a horizonal line at 0.9 sensitivity, with corresponding coordinate along the x-axis indicating false positive rate.

**Figure 3.**
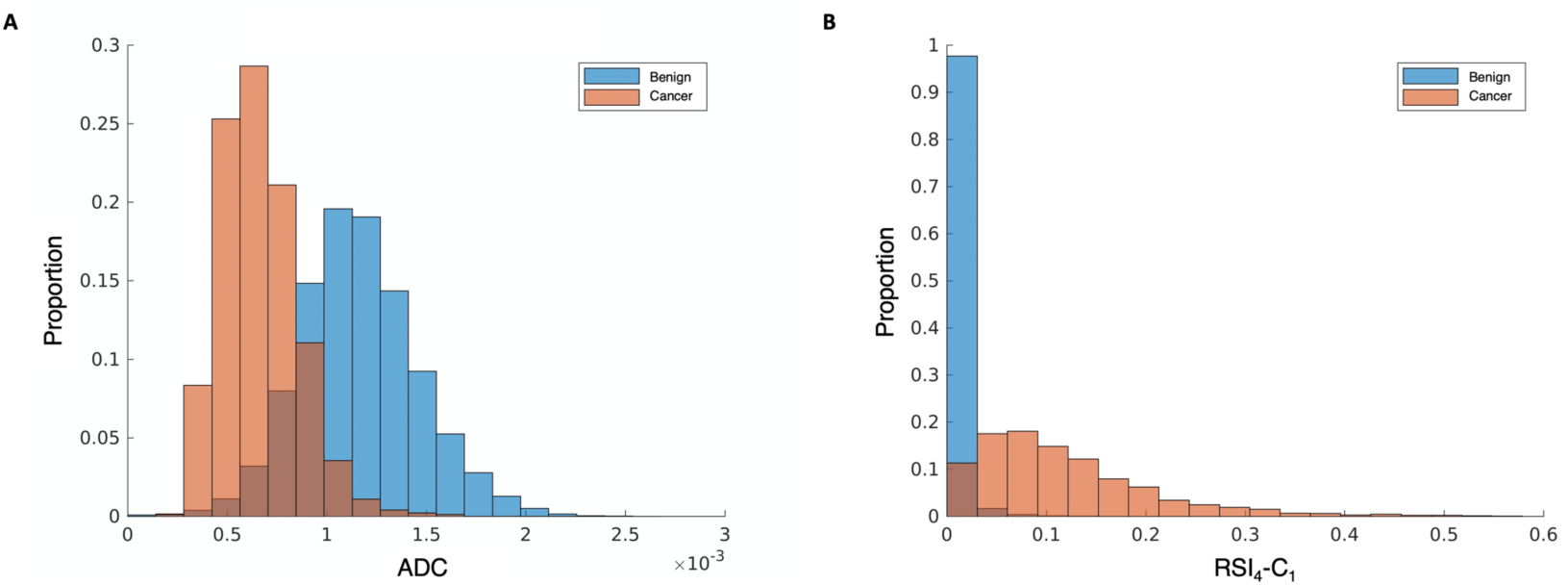
Normalized histograms of signal intensity for A) conventional ADC and B) RSI_4_-C_1._ Benign voxels are shown in blue and cancer voxels are in orange, with the overlapping regions in brown. RSI_4_-C_1_ has less overlap in the distribution of benign and cancer voxels compared to ADC.

**Figure 4.**
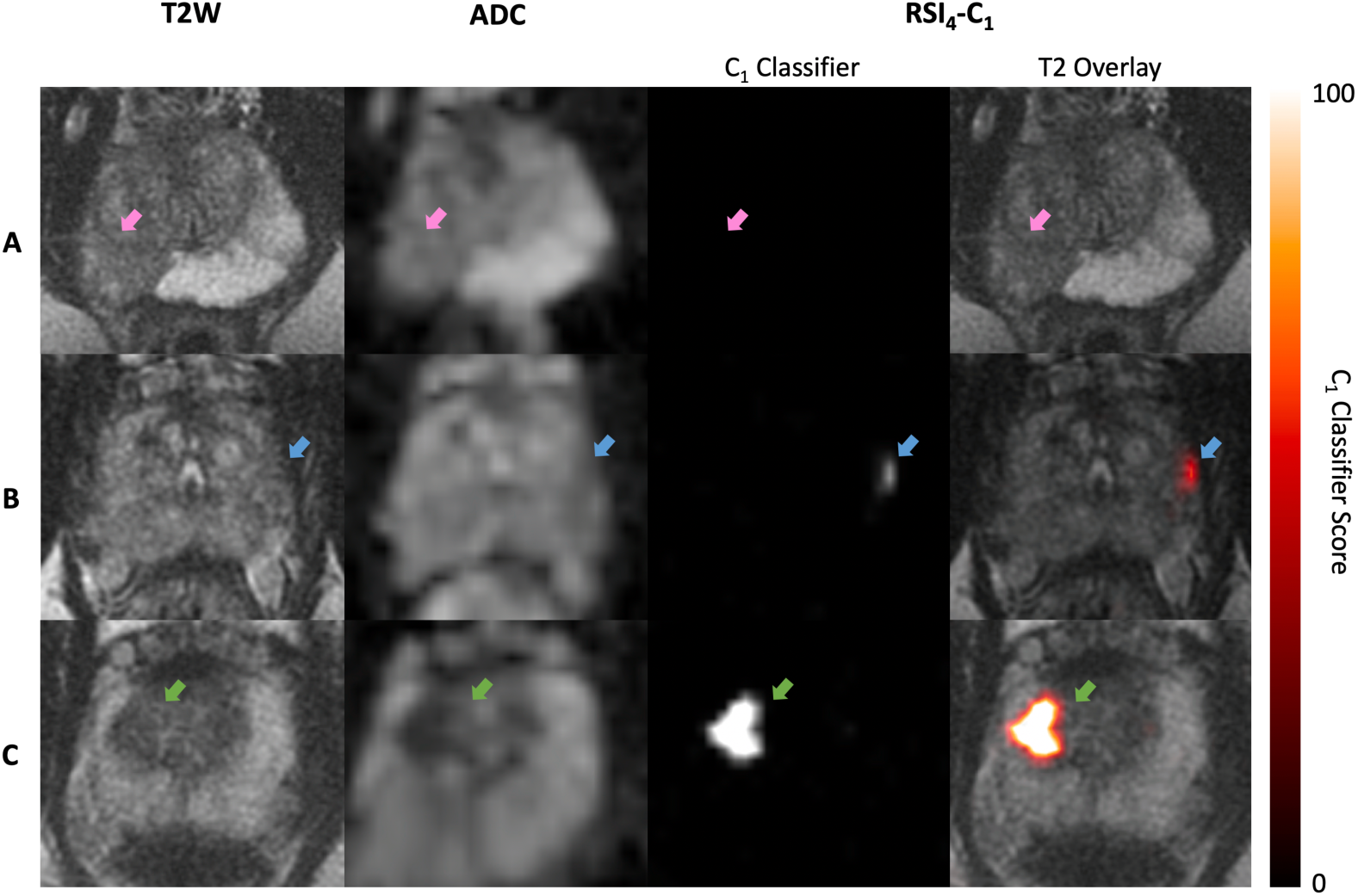
Representative axial images of T2-weighted MRI (T2W), conventional ADC, and logistic regression of RSI_4_-C_1_ (RSI_4_-C_1_ classifier) for 3 representative subjects. Subject A had a PI-RADS 5 lesion (pink arrow) on MRI, with two subsequent negative biopsies showing only acute and chronic inflammation. Subject B had a small PI-RADS 3 lesion (blue arrow) in the left peripheral zone; he underwent radical prostatectomy and was found to have Gleason 3+4 prostate cancer with focal ECE. Subject C had a PI-RADS 3 lesion (green arrow) in the right transition zone; he underwent prostatectomy and was found to have Gleason 4+3+5 prostate cancer. RSI_4_-C_1_ classifier maps readily highlight the cancers for subjects B and C. The RSI_4_-C_1_ classifier map for subject A has no false-positive voxels; it is shown on the same color scale as the maps for subjects B and C.

## Discussion

RSI_4_-C_1_ proved a superior voxel-level classifier for prostate cancer than conventional ADC, yielding significantly improved AUC and reduced false positives. When requiring 90% sensitivity for high-confidence cancer voxels in the cancer patients, conventional ADC performed poorly in control patients, falsely classifying approximately 1 in 5 benign voxels as cancerous (FPR_90_: 0.201, 95% CI 0.131-0.300). In contrast, for the same cancer sensitivity, RSI_4_-C_1_ gave far fewer false positives (FPR_90_: 0.033, 95% CI 0.009-0.083). This voxel-level classifier can be used to generate quantitative images that can be compared across subjects on the same scale and that highlight cancer with less noise (false positives) than the current imaging standard. These images may have utility in clinical applications such as MRI-guided prostate biopsy^20–22^ and targeted radiotherapy planning^23–25^.

To develop the RSI model, we selected voxels that were high confidence for either benign prostate or prostate cancer, using all available clinical and pathologic information. Surgical pathology was not available for all patients, but using consecutive patients and allowing heterogeneity in type of pathology specimen avoids the selection bias of a surgery-only group. High-confidence voxels were chosen to avoid introducing errors into the model. Because we used high-confidence voxels, we also expected high model performance, including high sensitivity for detecting these cancer voxels. The choice of FPR_90_ as a performance metric reflects this expectation: when requiring 90% sensitivity for high-confidence cancer voxels, a useful model will have a low false positive rate.

The discriminatory performance of RSI_4_-C_1_ relies on the RSI approach of separating the overall diffusion signal into compartments believed to correspond to restricted diffusion, hindered diffusion, free water, and rapid pseudo-diffusion. A prior study demonstrated improved characterization of diffusion signal within the normal prostate and prostate tumors with this four-compartment model, especially within the most diffusion-restricted compartment, C_1_^18^. By using this most restricted compartment, the vast majority of benign prostate tissue signal is suppressed, and output images have noticeably less noise (**Figure 4**) than conventional ADC maps. Prior studies have also investigated the performance and utility of advanced DWI techniques, including RSI, in prostate cancer detection and characterization^15,26–29^. However, many of the other studies conducted analysis at the lesion level rather than the voxel level. A voxel-wise classifier permits generation of cancer-detecting images, like those shown in Figure 4, and avoids the need to manually define lesions. Nevertheless, distinguishing malignant and benign lesions is an important clinical problem, as is distinguishing lower and higher-grade lesions. Future work will apply the voxel-level classifier output to lesion-level analyses in a larger dataset.

Conventional ADC was calculated in this study using the most widely utilized approach consistent with PI-RADS version 2.1^7^, the consensus standard for multi-parametric prostate MRI, which recommends that ADC maps be calculated with *b*-values less than or equal to 1000 s/mm^2^. Prior studies have reported increased conspicuity of prostate cancer when using *b*-values greater than 1000 s/mm^2 30–33^, and some centers—including ours—routinely acquire images with stronger diffusion weighting than that required by PI-RADS. However, the objective of the present work was to develop a quantitative, voxel-level classifier for prostate cancer. ADC is the clinical standard for quantitative diffusion MRI and so was chosen as the comparator to the quantitative model developed in this study. The diffusion weighted images, themselves, are typically interpreted qualitatively using subjective, patient-specific window/level settings. High *b*-value images do not lend themselves readily to a quantitative, voxel-level analysis without a model like the one described in the present work. Indeed, post-hoc analyses of the present dataset confirmed that no *b*-value yielded adequate voxel-level classification: the FPR_90_ for high *b*-value DWI (1000, 2000, and 3000 s/mm^2^) was well over 0.500 in each case, compared to 0.201 and 0.033 for conventional ADC and RSI_4_-C_1_, respectively.

There are some limitations to this study. We had a small sample size from a single scanner in order to take advantage of a specialized acquisition protocol, which may limit generalizability. This analysis does not compare the RSI_4_ model to other advanced DWI methods or investigate the potential added value of multiple echo times^14–16,26^; we plan to acquire data adequate for these comparisons for future analyses. As mentioned above, there was also heterogeneity in pathology type, which precluded voxel-level histopathology correlation but is reflective of real-life practice patterns. Relatively few transition zone cancers also precluded subset analysis of classifier performance by prostate zone. The overall excellent performance of our models may be partially attributed to use of majority PI-RADS category 4-5 cancers, which are already conspicuous for experienced radiologists. However, these lesions provided high-confidence training data.

In conclusion, our study demonstrates that RSI_4_-C_1_ yields a voxel-level classifier of prostate cancer that is superior to conventional ADC. RSI classifier images, with a lower false-positive rate, might be used to assist in accurate detection of prostate cancer. Future work will apply this RSI_4_-C_1_ classifier to a larger, independent dataset.

## Data Availability

The datasets used and analyzed during the current study are available from the corresponding author on reasonable request.

